# Household Vulnerability in the urban slums of Mumbai, India: Analysis of a large cross-sectional Survey

**DOI:** 10.1101/2023.07.20.23292961

**Authors:** Devika Deshmukh, Saurav Basu, Preeti Negandhi, Jyoti Sharma, Luigi D’Aquino, Vivek Singh, Mangesh Gadhari, Vaishali Venu, Rajeshwari Chandrasekar, Mangala Gomare, Sanjay Zodpey

**Affiliations:** UNICEF; PHFI; DFY; EHO, BMC

**Keywords:** Global Health, Urban health, Health vulnerability, Social vulnerability, residential vulnerability, occupational vulnerability, determinants of health, slums, India

## Abstract

**Background:** Improving equitable health outcomes needs a further understanding of the social, economic, political, and legal determinants that shape human health and well-being, especially in the poor and marginalized communities in urban slums. Vulnerability represents a group of adverse social determinants that put a household at a greater risk of falling ill.

The objective of this study was to determine the magnitude of health, residential, social, and occupational vulnerabilities amongst households in two urban slums in Mumbai, assess the sociodemographic factors associated with health vulnerability, and ascertain the linkage between health vulnerability and other vulnerabilities.

**Methods:** A cross-sectional survey was conducted from May to August 2021 in the urban slums of Mumbai. One Urban Primary Health Center area each in two wards (M/East and G/North) of Mumbai city mostly inhabited by people living in slums was purposively selected. A total of 15,796 households were included in the vulnerability assessment survey. Four kinds of vulnerability (health, social, residential, and occupational) indices were created based on survey responses.

**Results:** High residential vulnerability was estimated in 73.5%, (95% C.I. 72.8, 74.2), high social vulnerability in 67.9% (95% C.I. 67.2, 68.7), and high occupational vulnerability in 59.5%, (95% C.I. 58.7, 60.2) households. The presence of health vulnerability was observed in 39.6% (95% C.I. 38.8, 40.4) households. On adjusted analysis, social, residential, and occupational vulnerability were all statistically significant predictors of health vulnerability (p<0.001). The health vulnerability increased by 0.492 units for each unit increase in social vulnerability, 0.605 for each unit increase in residential vulnerability and 0.081 unit for each unit increase in occupational vulnerability.

**Conclusions:** Health vulnerability is present in nearly four out of ten households in the urban slums of Mumbai, while a majority of the households experience residential, social, and occupational vulnerability. Overcrowding and poor ventilation were nearly universal, with high burden of poor sanitation and hygiene.

## Introduction

India, in recent times, has been witnessing a rapid rise in urban populations. Individuals and families move to cities in search of lucrative jobs and better educational facilities; sometimes only with the hope for a better life ahead. This has led to unplanned urbanization in India, contributing to a growing slum population estimated at 35% of the total urban population (1,2). Slum dwellings are residential areas unfit for human habitation due to faulty design or construction, extreme overcrowding, crumbling infrastructure, faulty and narrow street design, with lack of ventilation, light, water, and/or sanitation facilities (3,4). People living in urban slums experience poverty, nutritional deprivation especially among women and children, low education, hazardous occupation, low-quality housing, unclean water supply, unregulated waste disposal, and the lack of open and safe recreational spaces, thereby adversely impacting their health (5–10). These populations also experience a significantly higher burden of communicable diseases and undernutrition compared to affluent populations (11–15). Furthermore, a combination of unhealthy diets, lack of avenues for regular exercise, and stressful social environments render the people living in the slums at high risk of non-communicable diseases (NCDs) including cardiovascular, diabetes, and hypertension (16–18). Vulnerability to disease and its associated complications are further accentuated in urban slums due to reduced accessibility, perceived quality, and underutilization of primary health care services (19–22). This lack of availability of essential health services significantly contributes towards increasing dependence of urban slum-dwelling population on the unregulated private health sector including unlicensed practitioners decreasing health equity and appropriateness of care (23–25).

The National Urban Health Mission (NUHM) was initiated as a submission of India’s landmark public health initiative – the National Health Mission (NHM) - in 2013, with the target of accelerating the progression toward universal health coverage and meeting essential health needs of the vast and neglected people living in urban slums by strengthening the urban public health system (26).

Relative deficiencies in social and health outcomes amongst slums previously were also observed in Mumbai, India which warrants differential allocation of resources by city administration (27). An assessment of the urban slums of Chandigarh city in North India ascertained that 36.3%, 51.2%, and 12.2% of the slums were categorized as highly, moderately, and low vulnerable, respectively (28). Consequently, the NUHM has recommended the vulnerability mapping and assessment activities for estimation of the residential, social, occupational, and health-related vulnerabilities of people living in urban slums towards conducting evidence-based urban health planning and implementation activities (29). However, to date, there has been no large-scale systematic analysis from empirical research in India applying NUHM recommended methodology that measures health vulnerabilities and their social determinants in these vulnerable demographics and informs the development of public health interventions towards a ‘health in all policies’ approach.

The primary healthcare systems in the developing world have been unable to adequately meet the essential health needs of the people living in the slums, especially related to their maternal and child health requirements (30). Moreover, during the Covid-19 pandemic, lack of primary health system preparedness and their suboptimal services, especially in the delivery of maternal and child health (MCH) and NCD treatment services, contributed to the further accentuation of the pre-existing health vulnerabilities amongst the people, especially living in slums (31).

The city of Mumbai, in India, is a mega-metropolitan with approximately 20.5 million population residing in the city with 52.5% of the population residing in slums (32). The National Family Health Survey (NFHS-5) (2019–20), a large-scale nationally representative cross-sectional survey observed that 18.6% children under 5 years of age were found to be underweight for their height in the Mumbai suburbia (33). Another cross-sectional analysis of 325 children between the ages of 10 and 18 months of mothers living in 20 urban slums in Western Mumbai reported that 76% of the children were anaemic, 31.2% had stunting, 25.1% were underweight, and 9.0% were wasted (34). In this context, it is important to identify, systematically measure and record the social deprivation and community health needs of this highly vulnerable population for the targeted development and application of evidence-based health interventions for mitigating their all-encompassing health risks.

This study was therefore conducted to determine the magnitude of health, residential, social, and occupational vulnerabilities amongst the people living in the households in two slums in Mumbai, assess the sociodemographic factors associated with health vulnerability, and ascertain the linkage between health and other vulnerabilities.

## Methods

Study design, duration, setting: A cross-sectional survey was conducted from May to August 2021 in urban slums of Mumbai. Two wards (M/East and G/North) in the city were purposively selected since they included two of the largest slums of the city. Subsequently, catchment areas of one of the fifteen urban primary health care (UPHC) facilities from the M/East and one of the nine facilities of G/North ward were again selected purposively due to consistently poor health indicators.

Sample size, strategy, and response: A census of all households in the two selected wards was taken for the study. A total of 26,496 households were mapped in the selected areas, but only 15,796 (59.6%) open households with eligible respondents were available for data collection (7674 - G/North, 8122 - M/East). Overall, 8994 (33.9%, N=26496) households in this area were closed or did not respond at the time of the visits, while 1706 (6.4%) households did not have any adult respondents and were ineligible for inclusion.

Vulnerability assessment in the urban slums was considered, as per the NUHM definition, as a group of adverse social determinants that put a household at a greater risk of falling ill [29]. The following four vulnerability domains, recognized as per the NUHM grouping, were operationally defined for this investigation:

- Health Vulnerability - level of vulnerability of a household towards ill-health and the extent of their inability to access appropriate healthcare services for achieving positive health outcomes.
- Residential Vulnerability - vulnerability associated with adverse housing settlement, lack of basic amenities such as WASH and/or environmental degradation.
- Occupational Vulnerability - vulnerability due to lack of regular employment and/or occupational hazards (such as rag picking, sex trade, mining, recycling waste collectors, construction workers, engaged in bidi making, matchbox making).
- Social Vulnerability - vulnerability due to social discrimination emerging out of an individual’s social status; experienced by women, children, elderly, transgenders, disabled, migrants, and minorities.

Questionnaire: The vulnerability assessment questionnaire was adapted from the Government of India’s validated vulnerability assessment and mapping questionnaire, and the National Multidimensional Poverty Index (29, 35). It consisted of four distinct modules to measure each vulnerability domain through a fixed set of questions: Residential Vulnerability (8 questions), Social Vulnerability (6 questions), Occupational Vulnerability (2 questions) and Health Vulnerability (8 questions) (Appendix). The questionnaire was subject to face validity by three public health experts. Pre-testing of the questionnaire was conducted to assess appropriateness in a different slum setting.

Each item response was coded as 0: Highest vulnerability, 1: Intermediate vulnerability, 2: No vulnerability. The residential, social, occupational, and health vulnerability scores were calculated using the cumulative score of all items within each vulnerability domain with each item having equal weightage. For each domain, a score that was equal to or above the median value was classified as low vulnerability, while those below the median value were classified as high vulnerability. A maximum possible score was considered as no vulnerability.

Data collection: To conduct the survey, field investigators were selected and trained at the outset. First, a map of the study areas was constructed and a transect walk was undertaken with community health workers to identify the distribution of the housing settlements within each selected ward. A census of all the households in the selected wards was included in this study. From each household, an adult respondent, preferably the head of the household, was interviewed face to face by a trained field volunteer using the questionnaire design. Each interview lasted for an average of 20 minutes, and each enumerator covered 25-30 households per day. Requisite COVID-19 appropriate behaviour was followed during the interview process to minimize the risk of infection transmission.

Data and Statistical Analysis: The data were electronically captured on Android smartphones using the Kobo toolbox application. This was uploaded in real-time on the internet-based servers. The backend team led by a data IT analyst screened the data for any missing values or any data discrepancy and provided necessary real-time feedback to the field investigators. All statistical analyses were performed using Stata version 15.1 (STATACorp, USA). Results were expressed in frequency and proportions for categorical variables, mean and standard deviation or median and interquartile range for continuous variables. Multivariate analysis through binary logistic regression was conducted to identify the factors independently associated with the presence of health vulnerability (dependent variables) after adjustment for other explanatory variables having significant statistical association on bivariate analysis. Finally, multiple linear regression was also carried out to predict health vulnerability based on the other vulnerability domain scores. A p < 0.05 was considered statistically significant.

Ethical Considerations: Administrative permission was taken from the Executive Health Officer (EHO), MCGM at the outset. Ethics clearance was also obtained from the Ethics Committee of Doctors For You prior to the commencement of data collection. Post obtaining ethics clearance, data collection was initiated in both slum areas. First, electronic consent was obtained from the head of the household for participation in the survey, given the COVID guidelines in place at the time of data collection. Those who consented were asked questions pertaining to vulnerability assessment. Anonymised data was entered real-time into the Kobo toolkit database, cleaned and used for analyses. No one other than the research team has access to the raw data, which has been safely stored in an e-folder maintained specifically for this study.

## Results

### Household characteristics

A total of 15796 households (8122 from M/East and 7674 from G/North ward) were included in the vulnerability assessment survey. Around 8994 households were closed or did not respond at the time of the survey and around 1706 households did not have any adult respondents and were ineligible for inclusion. The reasons for the non-response were, residents who had migrated to their villages during the first wave and had not returned, others declined due to their daily wage work, while others refused to take part in the survey without assigning any specific reason. The mean (SD) age of the respondents was 35 (12.2) years, including 32.6% male and 67.3% females. The sociodemographic characteristics of the participants are reported in Table A1.

The average household size was 4.8. Almost two-thirds (60.4%) of the households were nuclear families with a single earning member. Nearly 28% of the households had at least one child of school going age (6-15 years) not pursuing school education. Illiteracy among all adult male members was observed in nearly one in five (19.4%), and among all adult female members in nearly one in four (24.6%) households. Nearly half of the households (48%) were primarily earning their livelihood through daily wages.

### Vulnerability burden in urban slum households

Vulnerability assessment in the urban slums indicated the highest burden of residential vulnerability, followed sequentially by social, occupational, and health vulnerabilities. High residential vulnerability was estimated in 11,575 (73.5%, 95% C.I. 72.8, 74.2, n=15739) households. High social vulnerability was observed in 10,288 (67.9%, 95% C.I. 67.2, 68.7, n=15333) households. High occupational vulnerability was observed in 9,389 (59.5%, 95% C.I. 58.7, 60.2, n=15790) households. Presence of health vulnerability was observed in 6,253 (39.6%, 95% C.I. 38.8, 40.4, n=15786) households (Table 1). The M/East ward compared to the G/North had a higher burden of health, residential, social, and occupational vulnerabilities (p<0.001).

**Table 1.** Distribution of vulnerability in households of urban slums in Mumbai, India.

| Type of vulnerability | M/East (n=8120) |  | G/North (n=7675) |  | Total (N=15795)* |  |
| --- | --- | --- | --- | --- | --- | --- |
|  | n1 | % | n2 | % | n1+n2 | % |
| **Residential** |  |  |  |  |  |  |
| None | 16 | 0.2 | 58 | 0.8 | 74 | 0.5 |
| Low | 1431 | 17.6 | 2659 | 34.9 | 4090 | 26 |
| High | 6668 | 82.2 | 4907 | 64.4 | 11575 | 73.5 |
| **Social** |  |  |  |  |  |  |
| None | 0 | 0 | 0 | 0 | 0 | 0 |
| Low | 1401 | 17.7 | 3444 | 47.7 | 4845 | 32.1 |
| High | 6515 | 82.3 | 3773 | 52.3 | 10288 | 67.9 |
| **Occupational** |  |  |  |  |  |  |
| None | 1132 | 13.9 | 1394 | 18.2 | 2526 | 16.0 |
| Low | 1509 | 18.6 | 2366 | 30.8 | 3875 | 24.5 |
| High | 5479 | 67.5 | 3910 | 51.0 | 9389 | 59.5 |
| **Health** |  |  |  |  |  |  |
| None | 4239 | 52.2 | 5294 | 69.1 | 9533 | 60.4 |
| Low | 0 | 0 | 0 | 0 | 0 | 0 |
| High | 3881 | 47.8 | 2372 | 30.9 | 6253 | 39.6 |

\*Not adding to 100% due to missing data from certain households that precluded estimation of vulnerability types in those households

Health vulnerability indicators in the surveyed households included a distance of >2 km for nearest health facility (14.5%), death of a household member under 70 years of age within past 3 years (10.6%), chronic disability in any household member (14.1%), lack of visit by frontline/community health worker (15.8%), inappropriate health seeking behaviour (17.1%), missed routine immunization of children aged below 3 years (77.6%), absence of antenatal care during pregnancy (54.6%), and the absence of postnatal care (45.4%) in the past 12 months (Table 2).

**Table 2.** Distribution of Health Vulnerability components in Urban Slums, Mumbai.

| Item | M/East |  | G/North |  | Total |  |
| --- | --- | --- | --- | --- | --- | --- |
|  | n1 | % | n2 | % | N=n1+n2 | % |
| **1. Proximity to the Health facility** |  |  |  |  |  |  |
| Within the range of 2 km | 6239 | 76.8 | 7267 | 94.7 | 13506 | 85.5 |
| More than 2 kilometres | 1882 | 23.2 | 405 | 5.3 | 2287 | 14.5 |
| **2. Mortality status (age <70)** |  |  |  |  |  |  |
| Reported history of maternal death/child death/death due to TB, malaria or other infectious disease | 1264 | 15.6 | 403 | 5.3 | 1667 | 10.6 |
| No case of death of any household member below 70 years of age in last 3 years | 6857 | 84.4 | 7265 | 94.7 | 14122 | 89.4 |
| **3. Disability Status** |  |  |  |  |  |  |
| Member with chronic disability/debilitating illness like TB, AIDS, Cancer, Kidney failure | 1144 | 14.1 | 381 | 5 | 1525 | 9.7 |
| Household member with mild impairment but functional | 458 | 5.6 | 231 | 3 | 689 | 4.4 |
| No member with disability | 6519 | 80.3 | 7058 | 92 | 13577 | 86 |
| **4. Frequency of ANM visit** |  |  |  |  |  |  |
| Never | 1374 | 16.9 | 1063 | 13.9 | 2437 | 15.4 |
| Once in 3 months | 372 | 4.6 | 235 | 3.1 | 607 | 3.8 |
| Monthly | 6375 | 78.5 | 6374 | 83.1 | 12749 | 80.7 |
| **5. Preference for type of health facility\*** |  |  |  |  |  |  |
| Private Health Sector | 995 | 12.3 | 2485 | 32.4 | 3480 | 22 |
| Public Health Sector | 1552 | 19.1 | 779 | 10.2 | 2331 | 14.8 |
| Both (Government and Private) | 3365 | 41.4 | 3910 | 51 | 7275 | 46.1 |
| NGO | 238 | 2.9 | 33 | 0.4 | 271 | 1.7 |
| Chemist | 387 | 4.8 | 63 | 0.8 | 450 | 2.8 |
| Quacks/stores | 95 | 1.2 | 36 | 0.5 | 131 | 0.8 |
| Others | 124 | 1.5 | 77 | 1.0 | 201 | 1.3 |
| Do not take treatment in cases of illness | 1365 | 16.8 | 289 | 3.8 | 1654 | 10.5 |
| **6. ≥1 unimmunized child (1–3 years old) in household** |  |  |  |  |  |  |
| Household does not have any child | 4924 | 60.6 | 5549 | 72.4 | 10473 | 66.3 |
| Total number of households with eligible children | 3197 | 39.4 | 2118 | 27.8 | 5315 | 33.7 |
| Some children did not receive all doses of immunization | 883 | 10.9 | 305 | 4 | 1188 | 7.5 |
| All children received all doses of immunization | 2314 | 28.5 | 1813 | 23.6 | 4127 | 26.1 |
| **7. Antenatal care in pregnant women** |  |  |  |  |  |  |
| Household does not have any pregnant woman | 6957 | 85.7 | 6850 | 89.5 | 13807 | 87.5 |
| Total number of households with ANC women | 1164 | 14.3 | 806 | 10.5 | 1970 | 12.5 |
| Some pregnant women did not receive antenatal care | 705 | 8.7 | 188 | 2.5 | 893 | 5.7 |
| All pregnant women received antenatal care | 459 | 5.7 | 618 | 8.1 | 1077 | 6.8 |
| **8. Postnatal care (delivery within past 1 year)** |  |  |  |  |  |  |
| No eligible mothers in household | 6982 | 86.0 | 7095 | 92.7 | 14077 | 89.2 |
| Total number of households with eligible mothers | 1139 | 14.0 | 560 | 7.3 | 1699 | 10.8 |
| Some eligible mother did not get post-natal care | 731 | 9.0 | 197 | 2.6 | 928 | 5.9 |
| All eligible mother received post-natal care | 408 | 5.0 | 363 | 4.7 | 771 | 4.9 |

\* Multiple responses pooled to create item vulnerability score

Residential vulnerability indicators in the surveyed households included non-permanent house (88.6%), no separate room (79.7%), irregular drinking water (46%), hazardous location (12.4%), open defecation practice (21.3%), no water supply in toilet (13.8%), and no electricity connection (4.7%) (Table A2).

Social vulnerability indicators in the surveyed households included vulnerable families considered as single women or minor headed households (19%), lack of identity proof (4.2%), school dropout/no enrolment (21.6%), illiteracy in all adult males (19.1%), illiteracy in all adult females (24.6%) (Table A3). The proportion of daily wage earners in child or women headed households (68.8%) and single parent households (70.1%) was greater than in nuclear (59.8%) and joint family (55.1%) households, and this difference was statistically significant (p<0.001). One in five households were engaged in hazardous occupational practices (Table A4). The proportion of households with monthly income above Rs. 10,000.00 per month was significantly greater in the G/North (42.2%) compared to the M/East (32.8%) ward households (p<0.001) indicative of a comparatively improved economic status in G/North ward.

Health vulnerability (score) indicated positive correlation with residential, social vulnerability and occupational vulnerability with strength of correlation highest for residential, followed by social and then occupational (p<0.001). The strong correlation between health and the other vulnerability domains suggests the interconnectedness between these domains (Table 3).

**Table 3.**
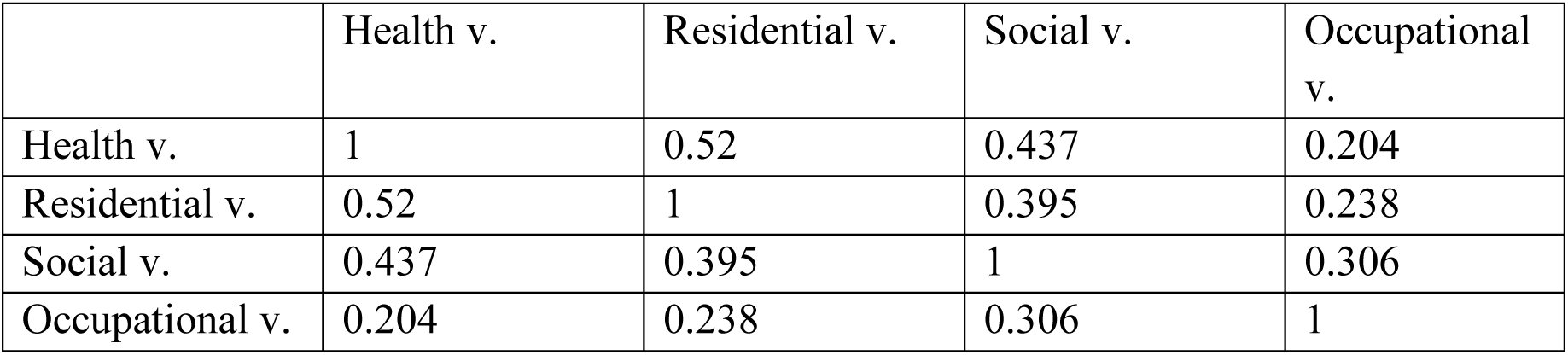
Correlation matrix of vulnerability domains in urban slums, Mumbai.

|  | Health v. | Residential v. | Social v. | Occupational v. |
| --- | --- | --- | --- | --- |
| Health v. | 1 | 0.52 | 0.437 | 0.204 |
| Residential v. | 0.52 | 1 | 0.395 | 0.238 |
| Social v. | 0.437 | 0.395 | 1 | 0.306 |
| Occupational v. | 0.204 | 0.238 | 0.306 | 1 |

On adjusted analysis, households with higher educational status of the female member, time since migration of greater than a year, non-hazardous nature of occupation were having significantly lower odds of the presence of health vulnerability(p<0.001). Religion of the household and highest educational status of the male member of the household were not significantly associated with health vulnerability status of the household. Households that had migrated within the past 1 year in comparison to those who were older migrants had 4.1 times higher odds of having health vulnerability. Households involved in hazardous occupation in comparison to non-hazardous occupational households had 2.9 times higher odds of health vulnerability (Table 4).

**Table 4.** Distribution of the factors associated with household health vulnerability.

| Characteristic | Total | Health vulnerability (present) | Adjusted odds | p-value |
| --- | --- | --- | --- | --- |
| Religion |  |  |  |  |
| Hindu | 7272 | 2926 (40.2) | 1 (Reference) | 0.077 |
| Muslim/Others | 8508 | 3323 (39.1) | 1.06 (0.99, 1.14) |  |
| Schooling male member |  |  |  |  |
| Up-to High School | 11712 | 4796 (40.8) | 0.93 (0.85, 1.02) | 0.147 |
| XIIth and above | 3945 | 1412 (35.8) | 1 (Reference) |  |
| Schooling female member |  |  |  |  |
| Up-to High School | 11748 | 4921 (41.9) | 1.22 (1.11, 1.34) | <0.001 |
| XIIth and above | 3496 | 1175 (33.6) | 1 (Reference) |  |
| Time since migration |  |  |  |  |
| <1 years | 1292 | 965 (74.7) | 4.12 (3.58, 4.74) | <0.001 |
| ≥1 year | 14494 | 5288 (36.5) | 1 (Reference) |  |
| Occupation |  |  |  |  |
| Hazardous | 3270 | 2090 (63.9) | 2.92 (2.68, 3.18) | <0.001 |
| Not Hazardous | 12159 | 4163 (33.3) | 1 (Reference) |  |

A multiple linear regression analysis was used to predict health vulnerability based on social, residential, and occupational vulnerability. A significant regression equation was found F (3, 15076) = 2558, p < 0.001 with an R2 of 0.337. The predicted household vulnerability is equal to 3.62 + 0.460 (Social Vulnerability) + 0.593 (Residential Vulnerability) + 0.081 (Occupational Vulnerability). The health vulnerability increased by 0.492 unit for each unit increase in social vulnerability, by 0.605 for each unit increase in residential vulnerability and by 0.081 unit for each unit increase in occupational vulnerability. Social, residential, and occupational vulnerability were all statistically significant predictors of health vulnerability (p<0.001) (Table 5).

**Table 5.** Multiple Linear Regression for health vulnerability predictors.

|  | Unstandardized<br>B | Coefficients<br>Std. Error | Standardized<br>Coefficient<br>Beta | t | Sig | 95%<br>CI<br>lower<br>bound | 95%<br>CI<br>upper<br>bound |
| --- | --- | --- | --- | --- | --- | --- | --- |
| (Constant) | 3.62 | 0.114 |  | 31.72 | <0.001 | 3.40 | 3.84 |
| Social V | 0.46 | 0.013 | 0.267 | 35.96 | <0.001 | 0.43 | 0.48 |
| Resident V | 0.59 | 0.011 | 0.411 | 56.40 | <0.001 | 0.57 | 0.61 |
| Occupation V | 0.081 | 0.021 | 0.027 | 3.89 | <0.001 | 0.04 | 0.12 |

Dependent variable: Health Vulnerability

## Discussion

Comprehensive primary healthcare services of high quality and affordability delivered through a patient-centric, community-based approach enabling preventive, promotive, curative, rehabilitative, and palliative care is an integral element of Universal Health Coverage (36). The present study observed significant challenges in accessing healthcare that corroborates prior evidence suggestive of major rural-urban discrepancies in access to care being comparable in urban slum populations [37]. The conceptual framework for health accessibility by Levesque et al (2013) situates healthcare accessibility in terms of multiple supply and demand side measures [38]. Challenges in primary healthcare accessibility in the households varied from difficulty in identifying health facilities among recent migrants to the lack of nearby facility, contributing to difficult access.

People living in densely populated urban slums in LMICs frequently experience the ‘crowding out’ phenomenon wherein uncontrolled queuing and congestion in public healthcare facilities restrict their utilization [39]. This study found that the lack of adequate staff in health facilities due to their diversion for pandemic management activities, and fear of contracting COVID-19 infection amongst patients were the major factors linked with disruption of continuum of care for antenatal and routine immunization services, corroborating previous evidence [40-42]. The failure to maintain linkages between the public health system and target beneficiaries in these resource limited settings emphasizes the need for greater sensitization and preparedness of health workers in maintaining existing health services.

A high prevalence of adverse social determinants of health in the surveyed urban slum population were observed in this study. Nearly one in five households were engaged in hazardous occupations that predispose the individual and household to a vicious cycle of poverty and ill health. Overlooking worker demands by application of power, apathy of employers, and systemic bias against workers contributes to persistent hazardous work conditions in India [43]. Educationally, more than one in four (28%) households had at least one child of school going age (6-15 years) that was either illiterate or dropout. Accentuation of this educational vulnerability in the marginalized population during the pandemic due to closure of schools and digital divide precluding utilization of online teaching is evident [44].

This study is one of the first in the country that was conducted for comprehensive vulnerability assessment in urban slums in India. The empirical findings indicate the interdependence of health with social, residential, and occupational vulnerabilities reflective of the need to improve public health holistically through a focus on social determinants across major domains. Such vulnerability assessments provide realistic data to policy makers and health programmers for the development of evidence-based policies, given their multi-sectoral coordination towards strengthening primary health care service delivery by identifying the more vulnerable groups and the associated social and economic barriers to improve healthcare accessibility. For instance, health systems should effectively track new migrants in slums by sensitization, monitoring and supervision through frontline health workers.

Vulnerability assessment exercises assist in providing detailed community profiles and household-level information on adverse social determinants that can be used to plan and respond with locally customized strategies. For instance, the households identified with occupational vulnerability due to hazardous work conditions can be subject to the ambit of occupational safety guidelines and screening for occupational exposure and disease by local officials. Similarly, female and child-headed households can be linked with all available social and health protection schemes especially during disasters and epidemics when their socioeconomic and health vulnerability is most accentuated due to loss of employment or wages. Recognition of the magnitude and predictors of specific vulnerability components can provide a useful resource for advocacy to local and municipal governments so as to advance the creation of an enabling environment sensitive to the needs of the vulnerable people living in the slums and promote optimal health seeking behavior through capacity strengthening and effective health communication.

The study strengths are the large sample size due to the census and estimation of multiple dimensions of vulnerability and their social determinants. However, as the methodology for the assessment included census several challenges were experienced during the survey. The areas surveyed comprised primarily of dense slums and compact housing and there was divergence in the method for mapping of the households compared to the local corporation’s assessment, as households with multiple entrances were marked as single households in this study but considered as separate households by the latter. Similarly, small-scale businesses/shops were not marked as households in this study unlike the corporation’s assessment. Nearly one third of the households were locked as many families had migrated during the pandemic; their information could not be captured during this survey. Further, this study could not capture individual attributes contributing to health, social and economic vulnerability such as addictions, specific occupational hazards, and nutritional status that should be evaluated in future studies.

In conclusion, this vulnerability assessment survey observed the presence of health vulnerability in nearly four in ten households in the urban slums of Mumbai. The residential vulnerability was present in nearly three in four households and was the strongest predictor of health vulnerability, while a majority of households had existing social and occupational vulnerabilities, suggestive of the interconnection of traditional adverse social determinants across the major vulnerability domains. Overcrowding and poor ventilation were nearly universal, with a high burden of poor sanitation and hygiene. Higher educational status of women was found to significantly reduce the health vulnerability of the entire household implying the need to strengthen efforts towards empowerment and educational efforts of women in urban slums. Improving equitable health outcomes needs regular tracking and measurement of vulnerabilities resulting from social, economic, political, and legal determinants that shape human health and well-being. This understanding of population vulnerabilities, especially from the poor and marginalized communities residing in informal settlements will ensure children’s right to life and thus support in achieving the Primary Healthcare and SDG goals.

## Data sharing statement

The anonymized dataset accompanying this submission would be made available on request to the corresponding author

## Acknowledgments

The authors acknowledge and thank Dr. Ajay Trakroo, Health Specialist, UNICEF New Delhi for his technical input and review and Dr Praween Kumar Agarwal, Research Officer, UNICEF, New Delhi for his critical input for data analysis.

## Competing interests

None. The author contributions are listed below

|  |  |
| --- | --- |
| Conceptualization | DD, RC, MGR, MG |
| Methodology | DD, SB |
| Validation | DD, SB, PN, JS, VV |
| Formal Analysis | SB, PN, JS |
| Investigation | DD, VV |
| Resources | DD, PN, SZ, MGR |
| Data Curation | DD, VV |
| Writing-Original Draft | SB, DD |
| Writing-Review and Editing | All authors |
| Visualization | DD, MG, SB, PN |
| Supervision | DD, RC, MGR |
| Project Administration | DD, MG, VS, LA, MGR |
| Funding acquisition | DD, MG, VS, RC, LA, PN, SZ |

\*MGR- Dr. Mangala Gomare, EHO, BMC

Patient and public involvement: This study did not involve patients. The findings of the survey are being shared with relevant stakeholders including the municipal corporation by the UNICEF to advocate and participate in improving health and reducing vulnerabilities through evidence informed programmatic action.

## SUPPLEMENTARY TABLES

### APPENDIX

**Table A1.**
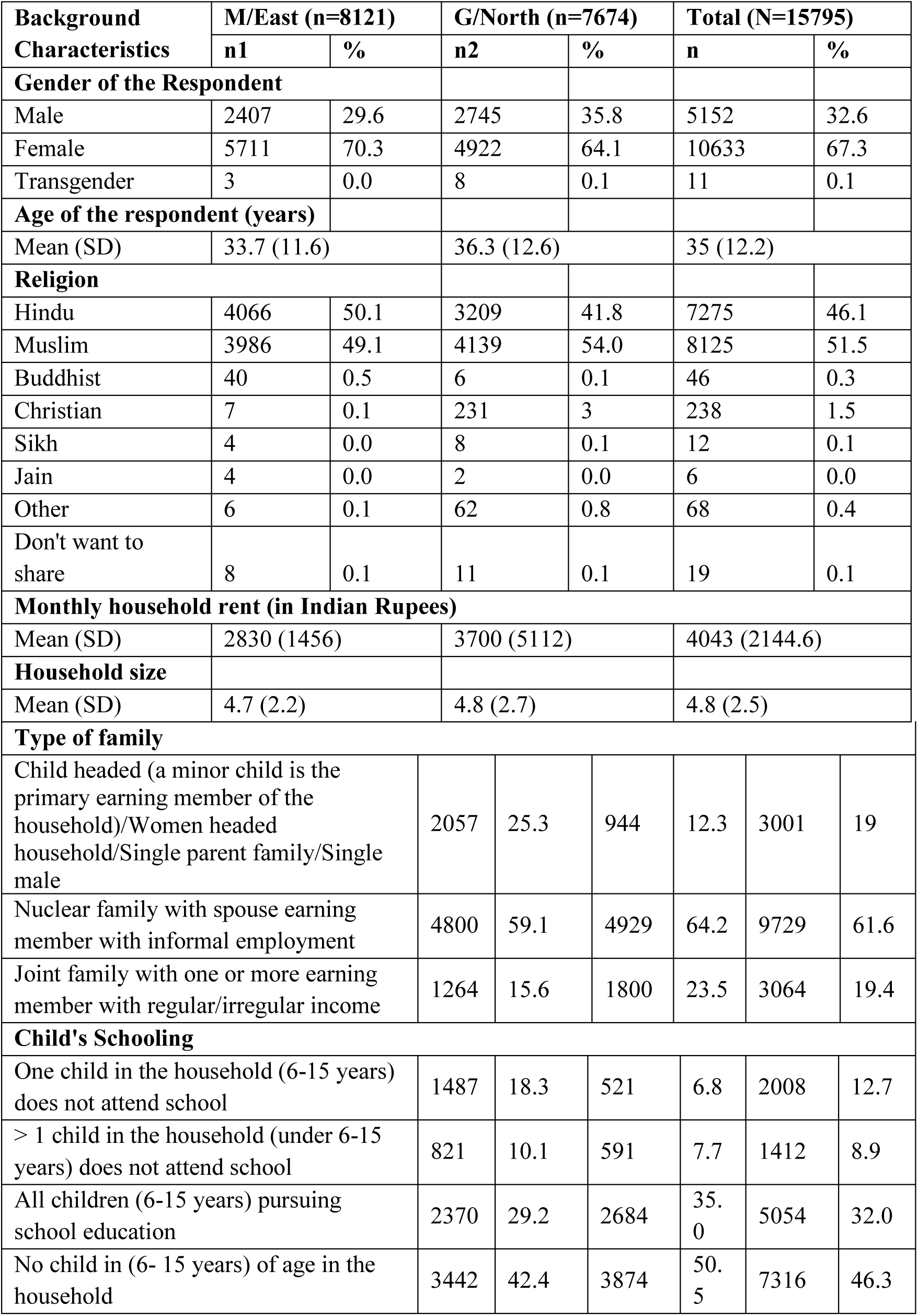
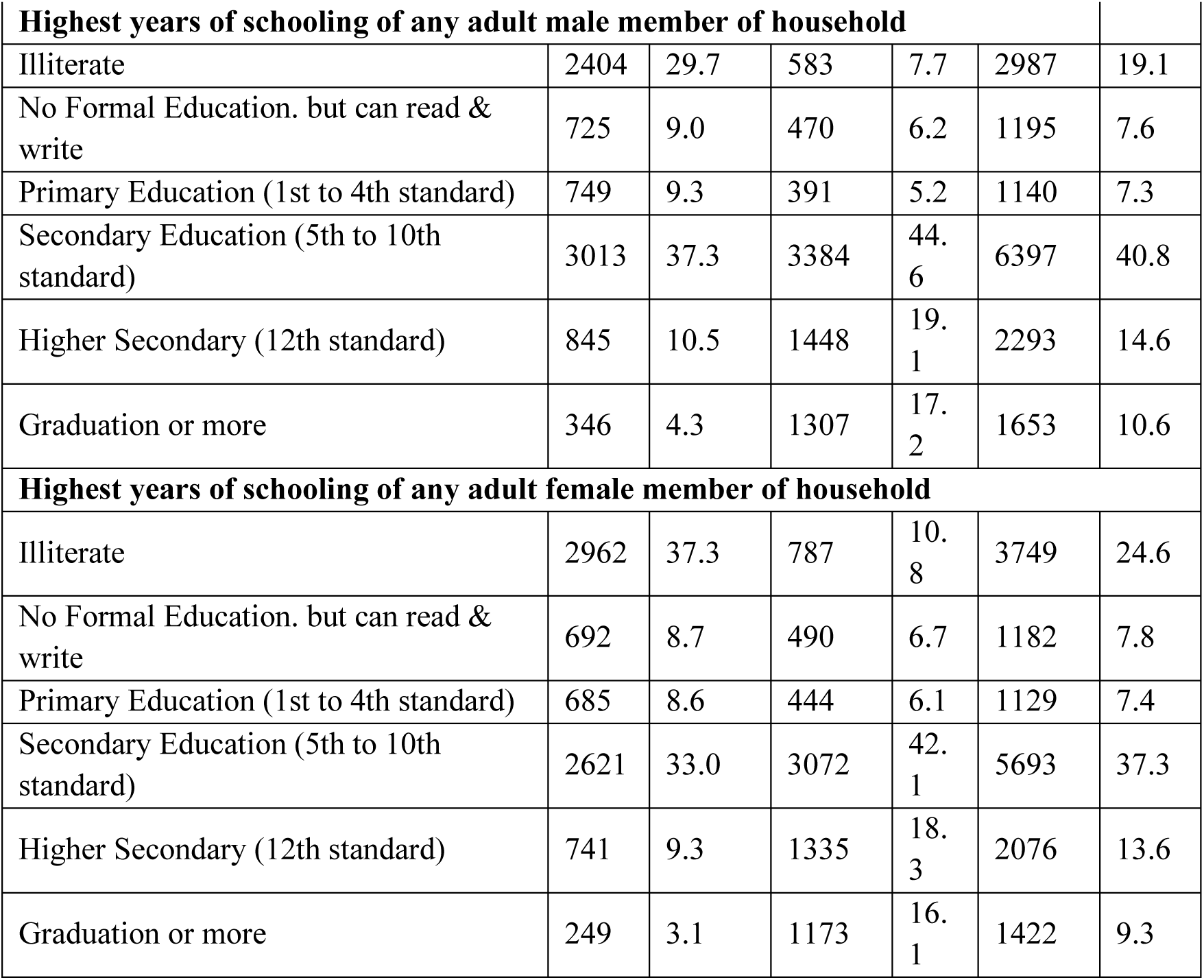
Sociodemographic characteristics of households in urban slums, Mumbai, 2021.

**Table A2.** Distribution of the Residential Vulnerability characteristics of households in Urban Slums, Mumbai, 2021.

| Characteristic | M/East |  | G/North |  | Total |  |
| --- | --- | --- | --- | --- | --- | --- |
|  | n1 | % | n2 | % | n | % |
| **1. Rooms in house** |  |  |  |  |  |  |
| No Separate room | 6257 | 77.0 | 6327 | 82.4 | 12584 | 79.7 |
| One bedroom | 1634 | 20.1 | 1234 | 16.1 | 2868 | 18.2 |
| Two and more bedrooms | 230 | 2.8 | 114 | 1.5 | 344 | 2.2 |
| **2. Slum characteristic** |  |  |  |  |  |  |
| Hazardous location besides dumping ground, polluted water, railway line or airport | 1710 | 21.1 | 248 | 3.2 | 1958 | 12.4 |
| Slum dwelling with high population density, poor ventilation, limited space | 4729 | 58.2 | 6648 | 86.6 | 11377 | 72 |
| Adequate ventilation and space | 1682 | 20.7 | 777 | 10.1 | 2459 | 15.6 |
| **3. Housing construction type** |  |  |  |  |  |  |
| Kutcha house with weak structure, no separate space for cooking, minimal ventilation | 1743 | 21.5 | 380 | 5 | 2123 | 13.4 |
| Semi- permanent but with mud/tin roof and non-cemented walls/brick walls with plastic or thatch roof | 5413 | 66.7 | 6463 | 84.2 | 11876 | 75.2 |
| Permanent structure, ventilation present, separate space for cooking | 965 | 11.9 | 832 | 10.8 | 1797 | 11.4 |
| **4. Toilet facility\*** |  |  |  |  |  |  |
| Open defecation (difficult access) | 2777 | 34.2 | 570 | 7.5 | 3347 | 21.3 |
| Have toilet (own / community), but open defecation | 1056 | 13 | 102 | 1.3 | 1158 | 7.4 |
| Use common/community toilet, do not have bath facilities (bath facility at home) | 2882 | 35.5 | 3516 | 46.1 | 6398 | 40.6 |
| Use common / community toilet, with bath facilities | 629 | 7.8 | 1440 | 18.9 | 2069 | 13.1 |
| Using own bathing and toilet facility | 772 | 9.5 | 2000 | 26.2 | 2772 | 17.6 |
| **5. Water supply in public toilet\*** |  |  |  |  |  |  |
| No | 233 | 6.6 | 945 | 18.9 | 1178 | 13.8 |
| Irregular (1 hour) | 544 | 15.5 | 344 | 6.9 | 888 | 10.4 |
| Irregular (1 -3 hours) | 142 | 4.0 | 86 | 1.7 | 228 | 2.7 |
| Irregular (3 + hours) | 228 | 6.5 | 211 | 4.2 | 439 | 5.2 |
| Yes, 24 hours | 2368 | 67.4 | 3414 | 68.3 | 5782 | 67.9 |
| **6. Public toilet cleaning frequency** |  |  |  |  |  |  |
| Daily | 2406 | 68.5 | 3624 | 72.5 | 6030 | 70.8 |
| Two or three days a week | 603 | 17.2 | 745 | 14.9 | 1348 | 15.8 |
| Weekly | 255 | 7.2 | 426 | 8.5 | 681 | 8 |
| Don't Know | 250 | 7.1 | 205 | 4.1 | 455 | 5.3 |
| **7. Public toilet cleaning personnel\*** |  |  |  |  |  |  |
| By private workers (agencies/ individual cleaners) | 1872 | 53.4 | 1860 | 39.3 | 3732 | 45.3 |
| By Municipal corporations / council / local body | 1396 | 39.8 | 2022 | 42.7 | 3418 | 41.5 |
| Don't Know | 238 | 6.8 | 854 | 18 | 1092 | 13.2 |
| **8. Drinking Water\*** |  |  |  |  |  |  |
| Regular (daily) | 6335 | 78.0 | 2190 | 28.6 | 8525 | 54.0 |
| Once a day | 909 | 11.2 | 5091 | 66.4 | 6000 | 38.0 |
| At least twice a day | 280 | 3.5 | 92 | 1.2 | 372 | 2.3 |
| Irregular (not daily) | 597 | 7.3 | 297 | 3.9 | 893 | 5.7 |
| Once a week | 28 | 0.3 | 12 | 0.2 | 40 | 0.3 |
| Twice a week (Less than once a week) | 22 | 0.3 | 16 | 0.2 | 38 | 0.2 |
| Three or more times a week | 22 | 0.2 | 6 | 0.1 | 27 | 0.2 |
| Purchase Water | 525 | 6.5 | 263 | 3.4 | 788 | 5.0 |
| **9. Type of public drainage facility\*** |  |  |  |  |  |  |
| Underground drainage | 2920 | 36.0 | 3249 | 42.3 | 6169 | 39.1 |
| Open with outlet | 2158 | 26.6 | 2229 | 29.0 | 4387 | 27.8 |
| Open without outlet | 708 | 8.7 | 1160 | 15.1 | 1868 | 11.8 |
| No drainage | 705 | 8.7 | 725 | 9.4 | 1430 | 9.1 |
| Don't know | 1628 | 20.0 | 310 | 4.0 | 1938 | 12.2 |
| **10. Electricity connection** |  |  |  |  |  |  |
| No electricity connections at all | 739 | 9.1 | 165 | 2.2 | 904 | 5.7 |
| Electricity without meter connection | 2496 | 30.7 | 666 | 8.7 | 3162 | 20.0 |
| Metered individual electricity connection | 4886 | 60.2 | 6841 | 89.2 | 11727 | 74.3 |

\* Multiple responses pooled to create item vulnerability score

**Table A3.** Distribution of the Social Vulnerability characteristics of households in Urban Slums.

| Item | M/East |  | G/North |  | Total |  |
| --- | --- | --- | --- | --- | --- | --- |
|  | n1 | % | n2 | % | n | % |
| **1. Type of Family** |  |  |  |  |  |  |
| Child headed / Women headed | 1005 | 12.4 | 411 | 5.4 | 1416 | 9 |
| Single parent family | 1051 | 12.9 | 533 | 6.9 | 1584 | 10 |
| Nuclear family with spouse earning member with informal employment | 4800 | 59.1 | 4929 | 64.2 | 9729 | 61.6 |
| Joint family with one or more earning member with regular income or irregular incomes | 1264 | 15.6 | 1800 | 23.5 | 3064 | 19.4 |
| **2. Identity Proof** |  |  |  |  |  |  |
| Have at least one but not all necessary documents (BPL card, Ration card, Voters ID, Aadhar) | 4268 | 52.6 | 2251 | 29.3 | 6519 | 41.3 |
| Have all the necessary documents | 3508 | 43.2 | 5107 | 66.6 | 8615 | 54.5 |
| Do not have any documents | 345 | 4.2 | 315 | 4.1 | 660 | 4.2 |
| **3. Type of Ration card\*** |  |  |  |  |  |  |
| Yellow (BPL) | 158 | 1.9 | 138 | 1.8 | 296 | 1.9 |
| Orange/Saffron (Priority Households) | 4663 | 57.4 | 5081 | 66.2 | 9744 | 61.7 |
| White (APL) | 112 | 1.4 | 159 | 2.1 | 271 | 1.7 |
| No Card | 3188 | 39.3 | 2295 | 29.9 | 5483 | 34.7 |
| **4. Child's Schooling** |  |  |  |  |  |  |
| One child in the household (6-15 years) does not attend school | 1487 | 18.3 | 521 | 6.8 | 2008 | 12.7 |
| > 1 child in the household (under 6-15 years) does not attend school | 821 | 10.1 | 591 | 7.7 | 1412 | 8.9 |
| All children (6-15 years) pursuing school | 2370 | 29.2 | 2684 | 35.0 | 5054 | 32.0 |
| Not applicable as no child in 6-15 years age group in the household | 3442 | 42.4 | 3874 | 50.5 | 7316 | 46.3 |
| **5. Highest years of schooling of any adult male member of household\*** |  |  |  |  |  |  |
| Illiterate | 2404 | 29.7 | 583 | 7.7 | 2987 | 19.1 |
| No Formal Education. but can read & write | 725 | 9.0 | 470 | 6.2 | 1195 | 7.6 |
| Primary Education (1st to 4th standard) | 749 | 9.3 | 391 | 5.2 | 1140 | 7.3 |
| Secondary Education (5th to 10th standard) | 3013 | 37.3 | 3384 | 44.6 | 6397 | 40.8 |
| Higher Secondary (12th standard) | 845 | 10.5 | 1448 | 19.1 | 2293 | 14.6 |
| Graduation or more | 346 | 4.3 | 1307 | 17.2 | 165<br>3 | 10.6 |
| **6. Highest years of schooling of any adult female member of a household\*** |  |  |  |  |  |  |
| Illiterate | 2962 | 37.3 | 787 | 10.8 | 374<br>9 | 24.6 |
| No Formal Education. but can read & write | 692 | 8.7 | 490 | 6.7 | 118<br>2 | 7.8 |
| Primary Education (1st to 4th standard) | 685 | 8.6 | 444 | 6.1 | 112<br>9 | 7.4 |
| Secondary Education (5th to 10th standard) | 2621 | 33.0 | 3072 | 42.1 | 569<br>3 | 37.3 |
| Higher Secondary (12th standard) | 741 | 9.3 | 1335 | 18.3 | 207<br>6 | 13.6 |
| Graduation or more | 249 | 3.1 | 1173 | 16.1 | 142<br>2 | 9.3 |

\*Multiple responses pooled to create item vulnerability score

**Table A4.**
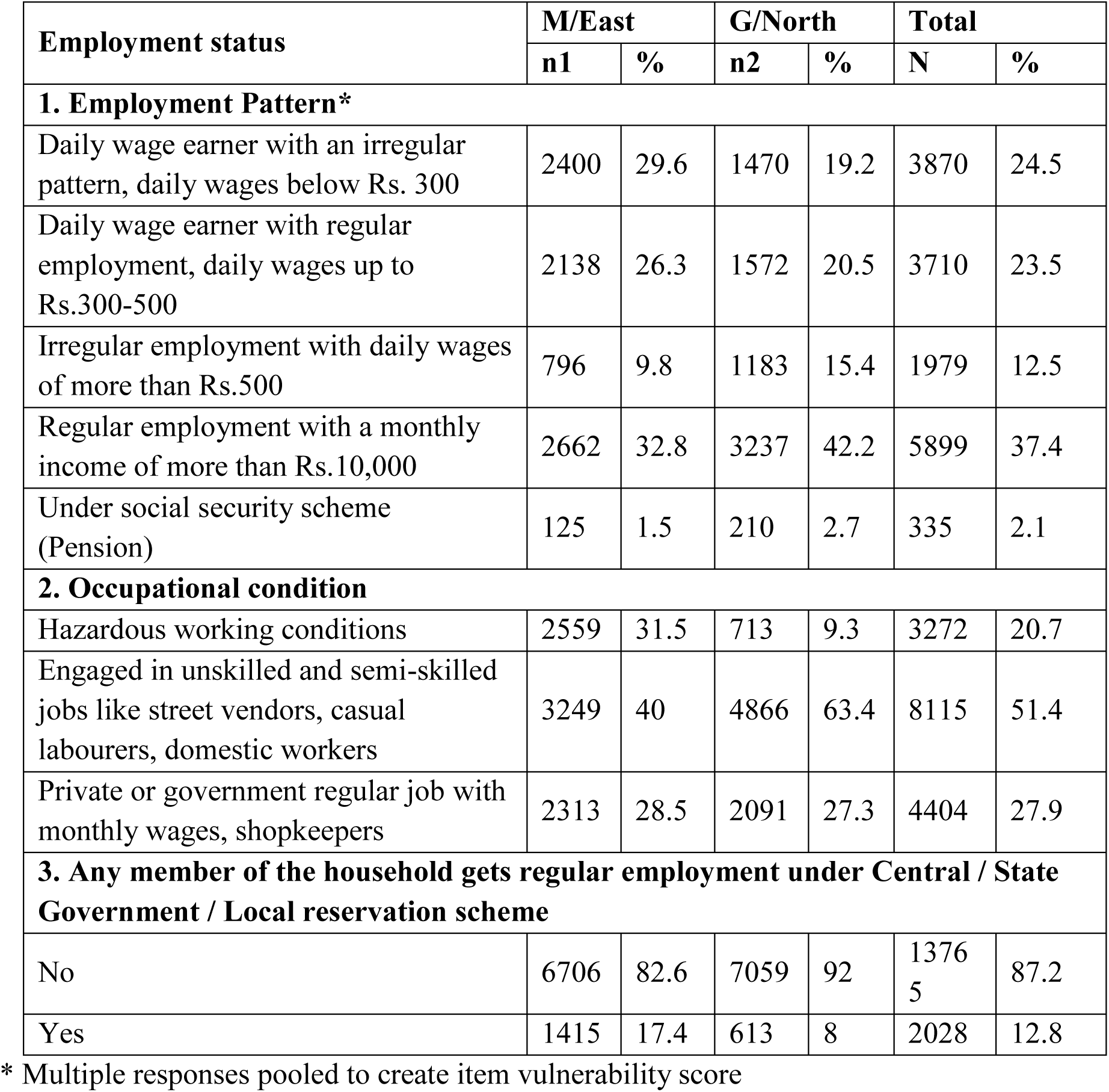
Distribution of the Occupational Vulnerability characteristics of households in Urban Slums, Mumbai.

| Employment status | M/East |  | G/North |  | Total |  |
| --- | --- | --- | --- | --- | --- | --- |
|  | n1 | % | n2 | % | N | % |
| **1. Employment Pattern\*** |  |  |  |  |  |  |
| Daily wage earner with an irregular pattern, daily wages below Rs. 300 | 2400 | 29.6 | 1470 | 19.2 | 3870 | 24.5 |
| Daily wage earner with regular employment, daily wages up to Rs.300-500 | 2138 | 26.3 | 1572 | 20.5 | 3710 | 23.5 |
| Irregular employment with daily wages of more than Rs.500 | 796 | 9.8 | 1183 | 15.4 | 1979 | 12.5 |
| Regular employment with a monthly income of more than Rs.10,000 | 2662 | 32.8 | 3237 | 42.2 | 5899 | 37.4 |
| Under social security scheme (Pension) | 125 | 1.5 | 210 | 2.7 | 335 | 2.1 |
| **2. Occupational condition** |  |  |  |  |  |  |
| Hazardous working conditions | 2559 | 31.5 | 713 | 9.3 | 3272 | 20.7 |
| Engaged in unskilled and semi-skilled jobs like street vendors, casual labourers, domestic workers | 3249 | 40 | 4866 | 63.4 | 8115 | 51.4 |
| Private or government regular job with monthly wages, shopkeepers | 2313 | 28.5 | 2091 | 27.3 | 4404 | 27.9 |
| **3. Any member of the household gets regular employment under Central / State Government / Local reservation scheme** |  |  |  |  |  |  |
| No | 6706 | 82.6 | 7059 | 92 | 13765 | 87.2 |
| Yes | 1415 | 17.4 | 613 | 8 | 2028 | 12.8 |

\* Multiple responses pooled to create item vulnerability score

